# Variations in Risk Factors across Different Periods of Stroke Recurrence

**DOI:** 10.1101/2023.12.14.23299993

**Authors:** Sang-Hun Lee, Jin-Man Jung, Jae-Chan Ryu, Moon-Ho Park

**Affiliations:** Department of Neurology, Korea University Ansan Hospital, Korea University College of Medicine, Ansan, Republic of Korea

**Keywords:** stroke recurrence, early recurrent stroke, late recurrent stroke

## Abstract

**Background and Purpose:** Accurately discerning periods of heightened risk of stroke recurrence and managing modifiable risk factors are essential for minimizing overall recurrence risk. This study identified differences in the timing of stroke recurrence based on risk factors and patient characteristics to develop strategies for reducing recurrence in clinical practice.

**Methods:** We retrospectively selected patients with ischemic stroke or transient ischemic attack at the Korea University Ansan Hospital Stroke Center between March 2014 and December 2021 using the prospective institutional database of the Korea University Stroke Registry. We collected demographic, clinical, and stroke data and categorized participants by recurrence timing (early within 3 months or late after 3 months). Using multinomial logistic regression analysis, we examined variables associated with early and late recurrent strokes.

**Results:** The analysis included 3,646 patients, of whom 255 experienced a recurrent stroke and 3,391 experienced their first stroke. Multinomial logistic regression analysis revealed significant associations between early recurrent stroke and diabetes mellitus (OR 1.98, 95% CI 1.25–3.15), other determined etiologies in TOAST classification (OR 3.00, 95% CI 1.37– 6.61), and white matter changes (OR 1.97, 95% CI 1.17–3.33). Late recurrent stroke showed a significant correlation with transient ischemic attack (TIA) (OR 2.95, 95% CI 1.52–5.71) and cerebral microbleeds (OR 2.22, 95% CI 1.32–3.75).

**Conclusion:** Our study emphasizes substantial differences in factors contributing to stroke recurrence based on timing. Managing the risk of recurrence in clinical practice necessitates accurate identification of heightened risk periods and rigorous control of modifiable risk factors.

## Introduction

Stroke is a disabling disease that causes high mortality rates and a significant decline in quality of life worldwide (1–3). Stroke exhibits a frequent recurrence compared to other diseases (4,5). Recurrent stroke causes more serious neurological damage, is more difficult to treat, and carries a higher risk of death, readmission, and long-term disability than the first stroke. Therefore, secondary prevention after the first stroke is critical to reducing stroke recurrence (6).

Understanding recurrent stroke is important. Previous studies have investigated important variables for stroke recurrence, including age, diabetes, smoking, arterial hypertension, hyperlipidemia, peripheral arterial disease, hypercoagulable states, depression, and the National Institutes of Health Stroke Scale (NIHSS) score status (7–10). The cumulative incidence of stroke recurrence in the first 5 years is 16–30% (11–13). However, the stroke recurrence rate varies significantly over time, particularly after the initial stroke. The timing of recurrence shows significant heterogeneity depending on the risk factors (3). The risk of recurrence ranged from 3.54 to 24.5% in the first year after the first stroke (14,15); the 2-year recurrence rate was approximately 30% (6); and in the following 5 years, it was in the range of 9.4% to 22.90% (16,17).

Precision in identifying the heightened risk of stroke recurrence and managing modifiable risk factors is crucial for minimizing the overall recurrence risk. The existing literature has overlooked early recurrence in the subacute phase, which occurs within 3 months after cerebral infarction. Consequently, our study aimed to address this research gap by identifying the risk factors and characteristics associated with early recurrence, particularly within the initial 3-month period. These findings provide valuable insights for clinical practice, aiding in the reduction of early recurrence rates.

## Methods

### Participants

This is a prospective institutional database from the Korea University Stroke Registry. The design of the database has been described in detail previously (18). Briefly, we retrospectively selected consecutive patients with ischemic stroke or transient ischemic attack (TIA) (diagnosed by a neurologist within 30 days of the event) who were admitted to the stroke center of Korea University Ansan Hospital between March 2014 and December 2021 due to neurological problems. Patients with hemorrhagic strokes were excluded from the study. (Figure 1) Stroke was defined as focal clinical signs of central nervous system dysfunction of vascular origin that lasted for at least 24 h according to the World Health Organization (Geneva) criteria. TIA was defined as losing cerebral or ocular function for less than 24 h. The recurrence of cerebral infarction was determined by neurological symptoms and ischemic changes detected on imaging tests.

**Figure 1.**
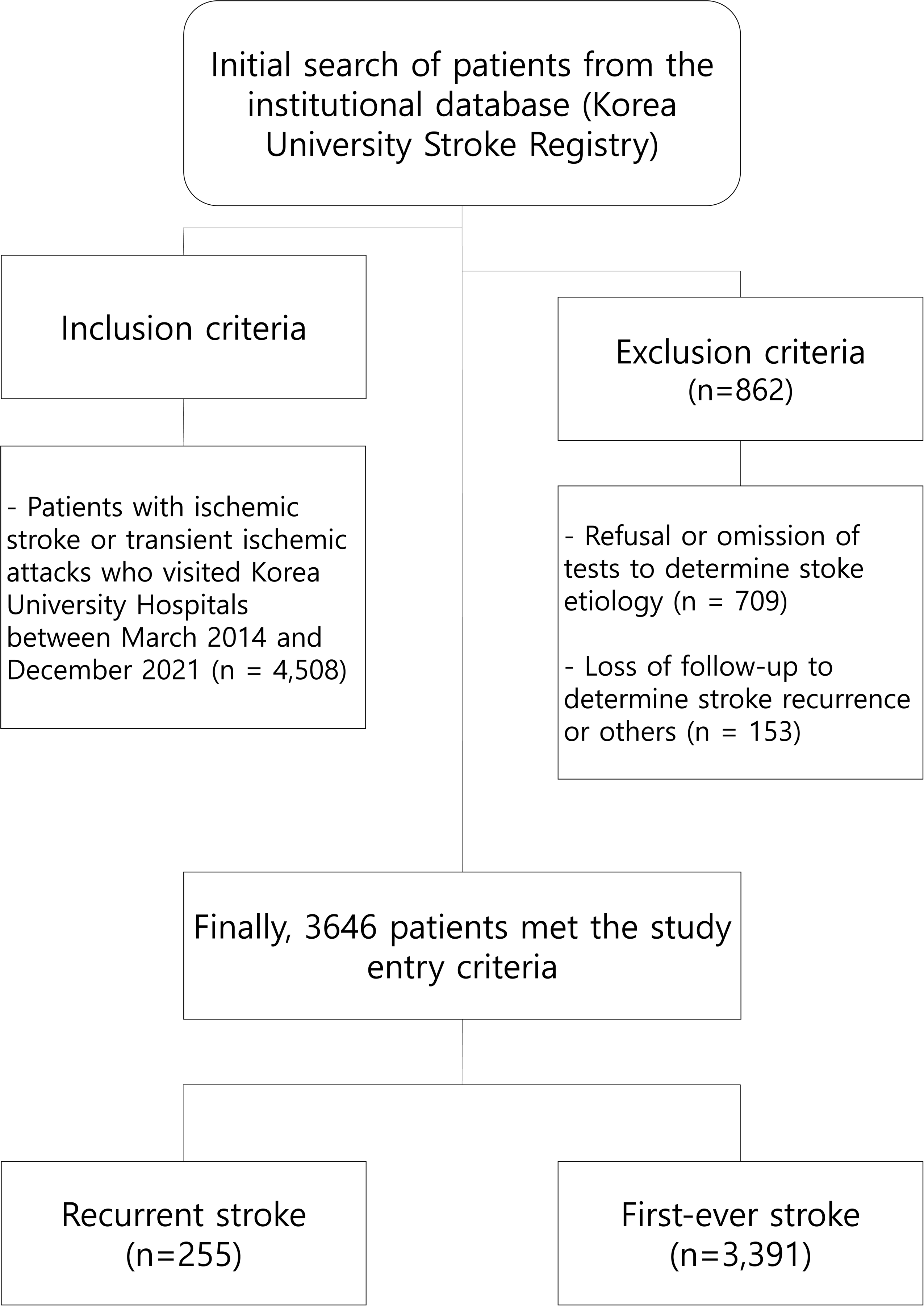
Flowchart of patient inclusion and exclusion criteria

Information on demographics, clinical evaluations, neurological examinations, stroke characteristics, and outcomes was obtained. The diagnosis required brain computed tomography (CT) and/or magnetic resonance imaging (MRI) to exclude hemorrhage and other causes of symptoms. Each patient required at least one vascular imaging scan, including conventional angiography, arterial imaging using MR angiography, CT angiography, or duplex ultrasound imaging. Standard systemic investigations were performed for every patient, including routine laboratory tests, chest radiography, and 12-lead electrocardiography. Routine laboratory tests included a complete blood count, electrolyte, glucose, renal function, liver function, lipid profile, and homocysteine levels. Transcranial Doppler, carotid duplex sonography, transthoracic echocardiography, transesophageal echocardiography, and 24-h Holter electrocardiography monitoring were performed in selected patients. This study was approved by the Institutional Review Board of Korea University Ansan Hospital (approval no. AS0213). Informed consent was not required owing to the retrospective design of the study.

### Clinical and Laboratory Assessment

For demographics and comorbidities, age at admission was categorized into three groups (<60, 60–74, and ≥75 years). Body mass index (BMI) was calculated as weight in kilograms divided by height in meters squared (kg/m^2^) and was categorized into four groups (underweight, <18.5; normal weight, 18.5–22.9; overweight, 23.0–24.9; obese, ≥25.0) by modified previous criteria (19). The weight and height required for BMI measurements were measured within 24 h of admission. The smoking status was classified as current smoker or nonsmoker. Hypertension was defined as a systolic blood pressure of at least 140 mmHg, a diastolic blood pressure of at least 90 mmHg, or the presence of hypertension when patients were previously diagnosed or were receiving antihypertensive medication. Diabetes mellitus was defined as a fasting serum glucose level of ≥126 mg/dL, a non-fasting serum glucose level of ≥200 mg/dL, a hemoglobin A1c level of ≥6.5%, or a history of insulin therapy and/or oral hypoglycemic drugs. Dyslipidemia was defined as a total cholesterol level of ≥200 mg/dL. Blood samples were obtained from all participants after at least 8 h of fasting on the morning of admission. Stroke is associated with abnormal biochemical parameters, including anemia (20), leukocytosis (21), elevated high-sensitivity C-reactive protein (CRP) levels (21), high homocysteine levels (22), and reduced kidney function (23). In this study, these biochemical cut-off points were used to define and categorize abnormal levels: anemia was defined as hemoglobin <12 g/dL in females and <13 g/dL in males; leukocytosis was defined as a white blood count of >12,000/μL; hyperhomocysteinemia ≥15 μmol/L; CRP elevation >2 mg/L; and abnormal kidney function as estimated glomerular filtration rate <60 mL/min per 1.73 m^2^.

Congestive heart failure was defined as a reduced left ventricular ejection fraction (<50%).

Atrial fibrillation (AF) was defined as persistent atrial arrhythmia with irregular R-R intervals and no clear repetitive P waves and was diagnosed with an electrocardiogram, 24-h Holter, or continuous electrocardiogram monitoring during hospitalization.

White matter changes (WMCs) in the hemisphere contralateral to the area affected by the acute stroke were assessed using the proposed visual rating scale (24,25). If the WMCs were higher on one side, the rating was based on the less involved or uninvolved side with the assumption of symmetry. In this study, the WMCs rating was modified using a dichotomous method (WMCs, grade 3 *vs.* no WMCS, grades 1–2). Cerebral microbleeds (CMBs) were assessed as rounded or ovoid hypointense lesions on a T2-GRE-weighted sequence of MRI (26). CMBs measured 10 mm or less in diameter and were surrounded by brain parenchyma over at least half the circumference of the lesion. The severity of neurological deficits at admission was rated using the National Institutes of Health Stroke Scale (NIHSS) score (27) on the day of admission, defined as poor initial NIHSS (NIHSS score ≥5), which was evaluated by the certified neurologists who were blind to this study.

For subtypes of stroke, ischemic stroke was classified according to the Trial of Org 10172 in the Acute Stroke Treatment (TOAST) classification system (28): large artery disease (LAD), cardioembolism (CE), small vessel occlusion (SVO), stroke of other determined etiology (OD), and stroke of undetermined etiology (UD). The UD category included three heterogeneous groups: “two or more causes,” “negative evaluation,” and “incomplete evaluation.” Recurrent stroke was defined as a new neurological deficit or deterioration of the previous deficit and fit the definitions for ischemic stroke, or TIA. Recurrent stroke was classified according to the recurrent interval as early recurrent stroke (≤3 months) or late recurrent stroke (>3 months). The extent of the diagnostic workup, stroke, and TIA were determined primarily by the stroke neurologists in charge of the patients, and the types of stroke and TIA were confirmed at a monthly stroke registry meeting.

### Statistical Analysis

All participants were categorized according to stroke recurrence (recurrent stroke group and first-ever stroke group), and the recurrent stroke group was further subcategorized into early and late recurrent stroke groups. Categorical variables are described in terms of frequencies and percentages. Differences in characteristics among groups were analyzed using Pearson’s chi-squared (*χ*^2^) test or Fisher’s exact test, as applicable. A pairwise *z*-test with the Bonferroni correction was used to determine the significance of the contribution of each subgroup of variables.

To identify potential predictors of recurrent stroke, we selected variables with *P* <0.2 in a bivariable analysis. We used a modified Nightingale-Rose diagram to show the distribution of potential predictors of recurrent stroke. These potential variables of recurrent stroke were used to generate a predictive model for each recurrent stroke by multivariable binary and multinomial logistic regression models using bootstrapping methods with a statistically significant association at *P* <0.05 compared to those of first-ever stroke. Goodness-to-fit and pseudo-R square statistics were used to evaluate the model fit and predictive strength. The final models were used to calculate the adjusted odds ratios (ORs) and 95% confidence intervals (CIs).

Statistical significance was declared when the two-tailed *P*-value was <0.05. Statistical analyses were performed using R version 4.2.1 software (R Foundation for Statistical Computing, Vienna, Austria) and SPSS (version 20.0; IBM SPSS, Chicago, IL, USA).

## Results

Between March 2014 and December 2021, 4,508 individuals with ischemic stroke or transient ischemic attack (TIA) were admitted to the Stroke Center at Korea University Ansan Hospital. Of these, 862 patients were excluded from the final analysis, comprising 709 patients who refused the omission of tests for the stroke registry and 153 patients who were lost during the evaluation period for stroke recurrence. Consequently, the final analysis included 3,646 patients, of whom 255 experienced recurrent strokes and 3,391 had their first stroke. (Figure 1)

### Differential Clinical Characteristics in Recurrent Stroke Versus First-Ever

Table 1 outlines the demographic and clinical features of the participants according to the occurrence of stroke recurrence. The recurrent stroke patient group had a significantly higher proportion of men than the first-ever stroke group (71.4% vs. 60.4%, p = 0.001). Variations in BMI across categories, such as normal, underweight, overweight, and obese, showed that patients with the underweight category (BMI 18.5–22.9) showed a significantly higher rate of recurrent stroke (9.1% vs. 4.4%, p = 0.005).

**Table 1.**
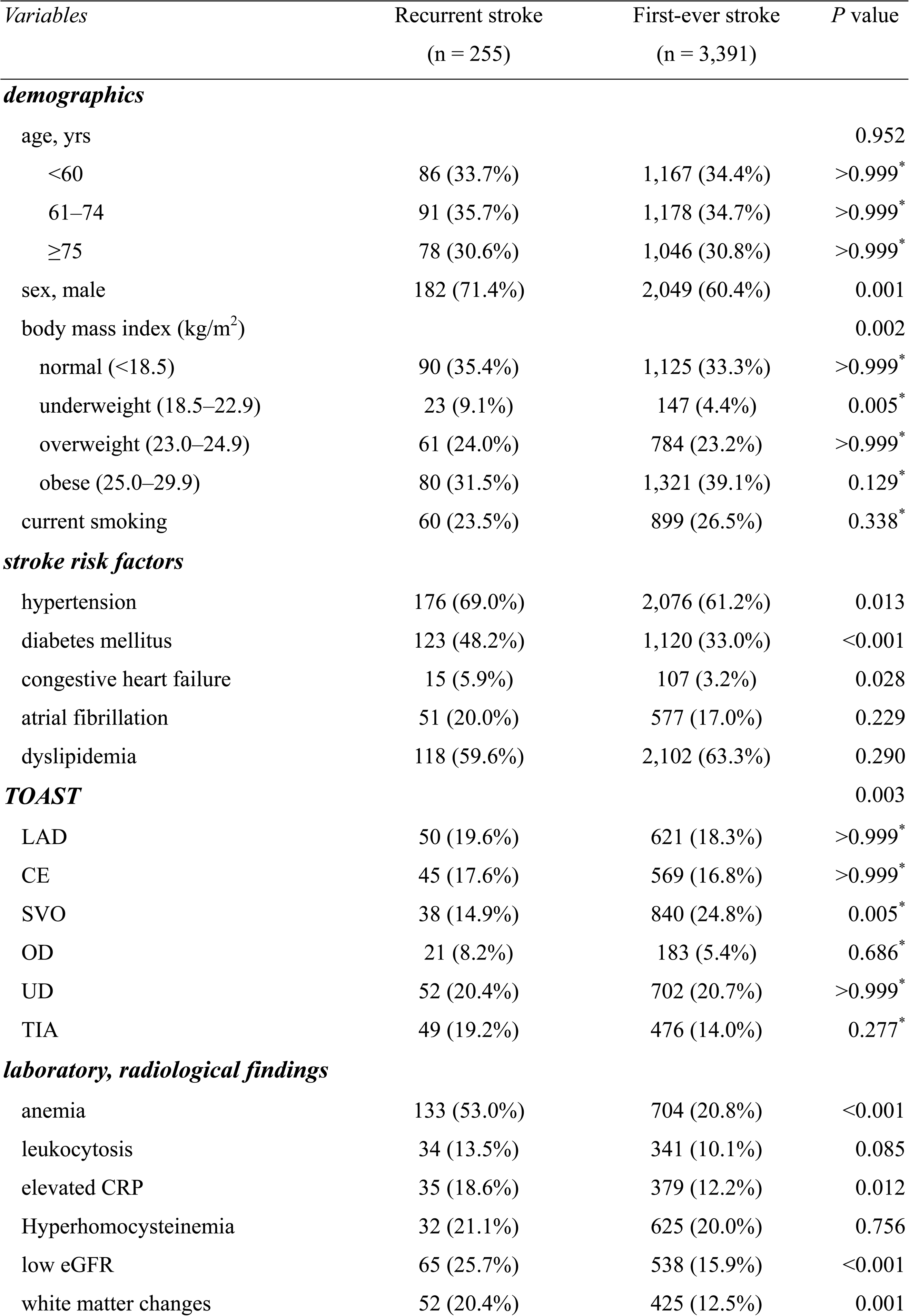
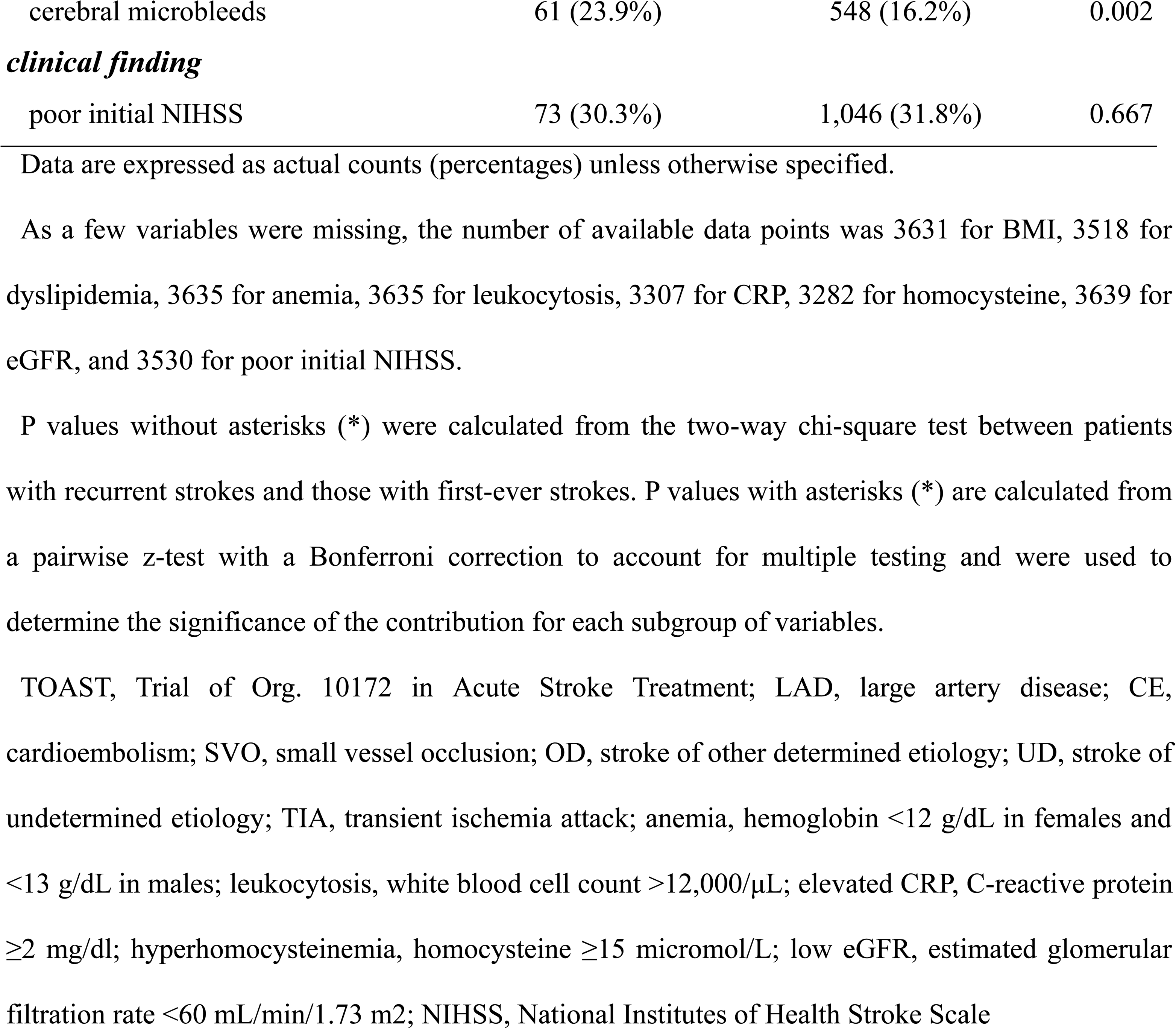
Clinical characteristics according to stroke recurrence.

Concerning stroke risk factors, higher recurrence rates were observed in individuals with hypertension, diabetes mellitus, and congestive heart failure. Stratifying participants based on stroke subtype according to the TOAST (Trial of Org. 10172 in Acute Stroke Treatment) classification revealed a significantly lower recurrence rate in cases of small vessel occlusion (SVO) (14.9% vs. 24.8%, p = 0.005).

Laboratory findings indicated a higher prevalence of anemia, elevated CRP levels exceeding 2 mg/dL, and a low estimated glomerular filtration rate (eGFR) in recurrent cerebral infarctions. In terms of radiological findings, white matter changes and cerebral microbleeds were associated with recurrent strokes.

### Differences in Clinical Characteristics among Early Recurrent Stroke, Rate Recurrent Stroke, and First-Ever Stroke

Figure 2. summarizes the analysis of the differences between the initial recurrent stroke, recurrent stroke rate, and first stroke using a diagram. For variables such as male sex, diabetes mellitus, TOAST classification, anemia, and cerebral microbleeds, patients with early and late recurrent stroke showed similar patterns of significant differences when compared with patients with first-ever stroke.

**Figure 2.**
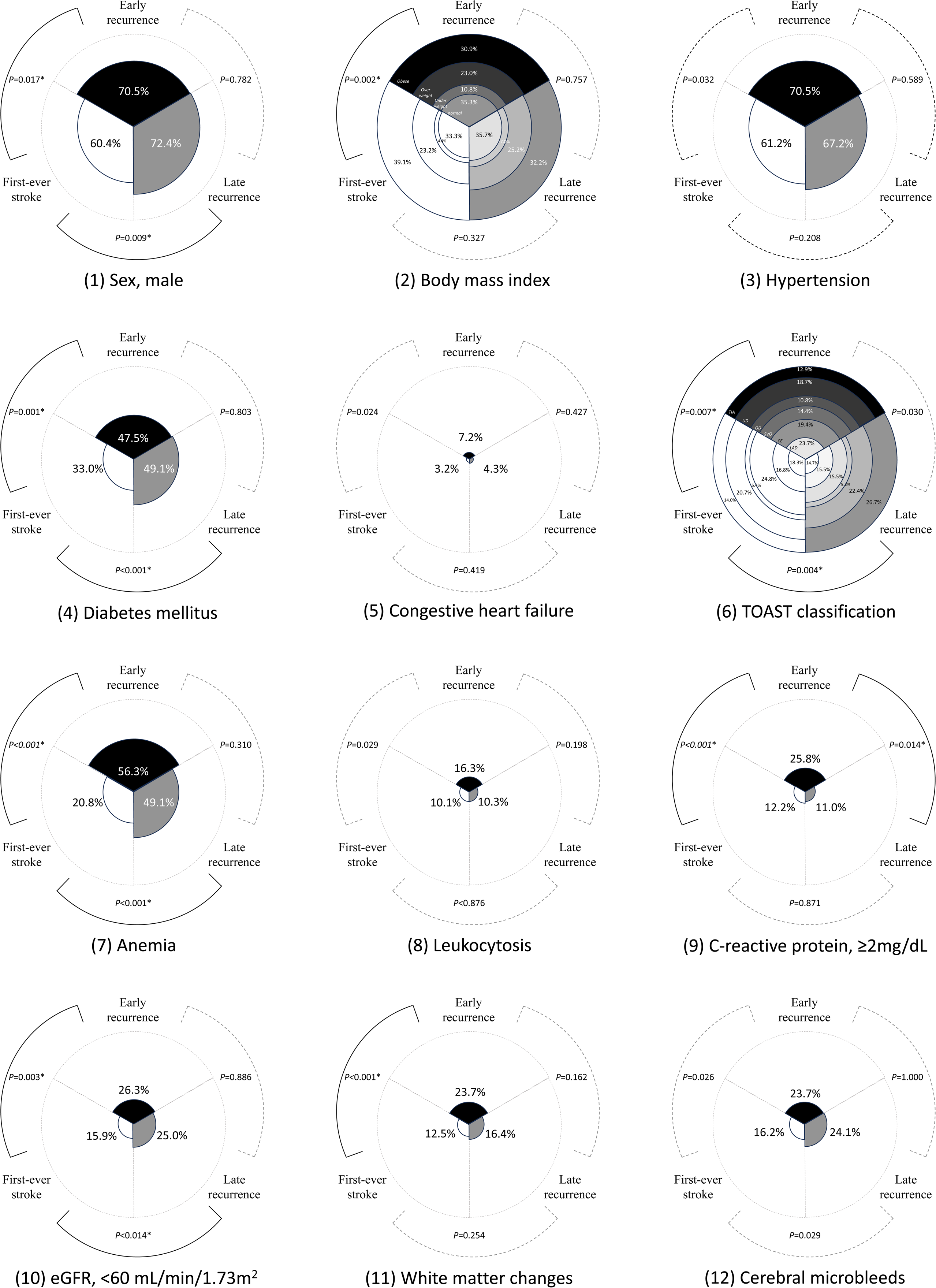
Major differences in clinical characteristics among early recurrent stroke, late recurrent stroke, and first-ever stroke Black (▪) circular sector indicates early recurrent stroke, gray(▪) indicates late recurrent stroke, and white (□) indicates first-ever stroke. These variables were selected, which had significant distribution in a bivariable analysis (P value <0.2 in Table 1). P values were calculated using the chi-square test with Bonferroni correction to determine the significant distribution among early recurrent stroke, late recurrent stroke, and first-ever stroke. P values with asterisks (*) and black solid lines indicate statistical significance. P values without asterisks and gray dotted lines indicate no statistical significance.

Regarding BMI (p = 0.002), congestive heart failure (p = 0.024), leukocytosis (p = 0.029), and white matter changes (p <0.001), significant differences were observed in comparison to first-ever stroke patients only in the early recurrent stroke group. However, regarding dyslipidemia (p = 0.047), significant differences in patients with first-ever stroke were observed only in the late recurrent stroke group.

A comparison of early and late recurrent strokes showed significant differences in TOAST classification and CRP levels. This is expressed in detail in Supplementary Table 1.

### Multivariable Logistic Regression Analysis of Predictive Factors for Early and Late Recurrent Stroke

A multinomial logistic regression analysis was used to analyze the early and late recurrent stroke variables. (Figure 3) For overall recurrent stroke, male sex (OR 2.05, 95% CI 1.45–2.88), hypertension (OR 1.54, 95% CI 1.06–2.22), diabetes mellitus (OR 1.59, 95% CI 1.15–2.18), other determined etiologies in TOAST classification (OR 1.98, 95% CI 1.04–3.78), TIA (OR 2.01, 95% CI 1.18–3.43), anemia (OR 3.96, 95% CI 2.81–5.57), white matter changes (OR 1.62, 95% CI 1.08–2.44), and cerebral microbleeds (OR 1.88, 95% CI 1.29– 2.72) showed significant results.

**Figure 3.**
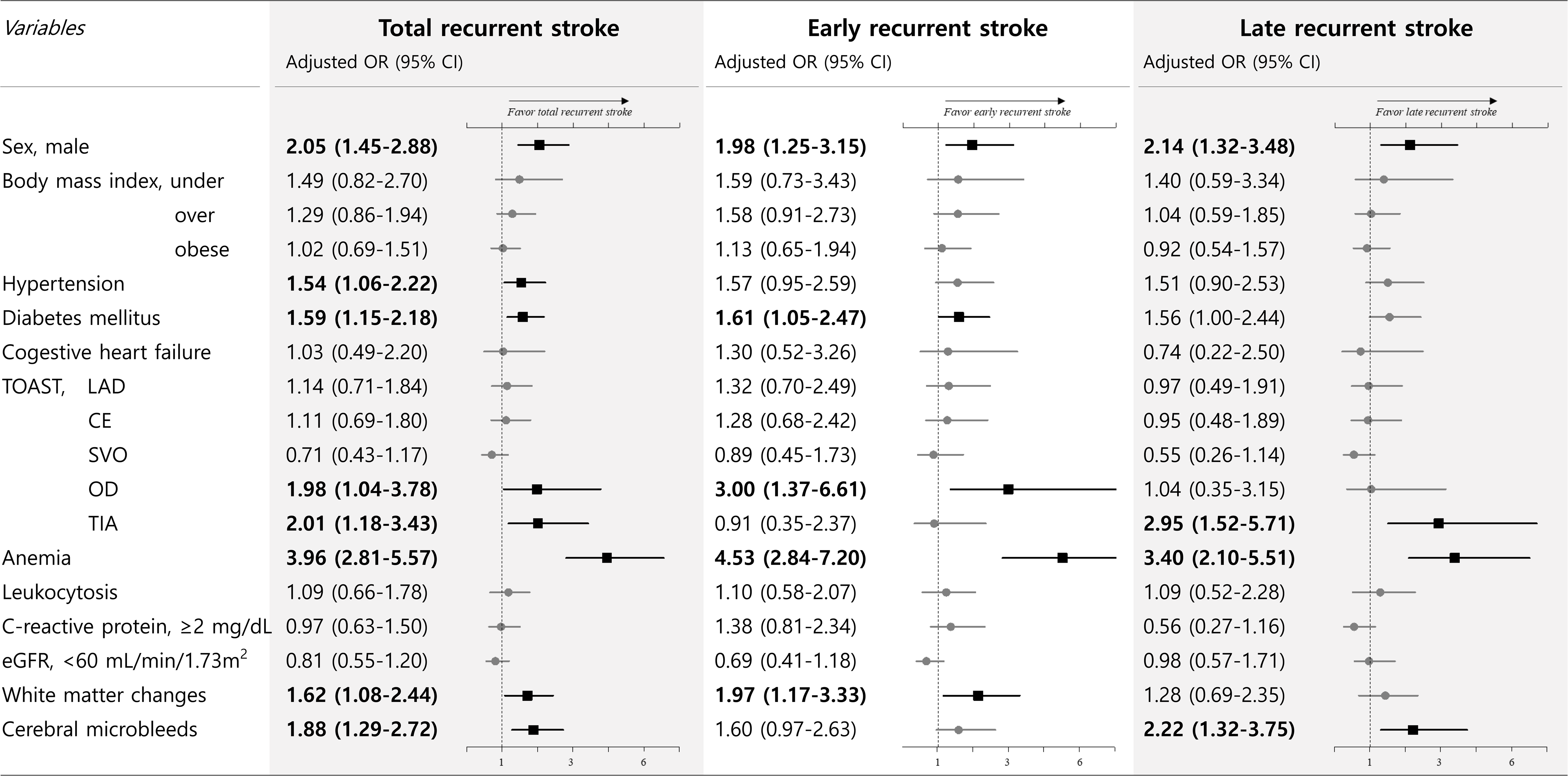
Predictive factors for each recurrent stroke by multivariable logistic regression analysis Values were expressed as odds ratios (95% CI). Polygons and lines in the plot are expressed as odds ratios (ORs) with a 95% CI. These odds ratios were adjusted for all covariates with a P value <0.2, as observed in the univariate analysis. Binary logistic regression analysis was used to analyze the variables associated with total recurrent stroke. A multinomial logistic regression analysis was used to analyze the early and late recurrent stroke variables. Values in bold font, black squares, and black lines indicate statistical significance (P<0.05). In contrast, values in normal font, gray circles, and gray lines indicate no statistical significance. The results were adjusted for sex, body mass index, hypertension, diabetes mellitus, congestive heart failure, TOAST, anemia, leukocytosis, CRP, ≥2 mg/dL, white matter changes, and cerebral microbleeds. TOAST, Trial of Org. 10172 in Acute Stroke Treatment; LAD, large artery disease; CE, cardioembolism; SVO, small vessel occlusion; OD, stroke of other determined etiology; UD, stroke of undetermined etiology; TIA, transient ischemia attack; anemia, hemoglobin <12 g/dL in females and <13 g/dL in males; leukocytosis, white blood cell count >12,000/μL; elevated CRP, C-reactive protein ≥2 mg/dl; hyperhomocysteinemia, homocysteine ≥15 micromol/L; low eGFR, estimated glomerular filtration rate <60 mL/min/1.73 m2; NIHSS, National Institutes of Health Stroke Scale

In the context of early recurrent stroke, significant associations were observed for diabetes mellitus (OR 1.98, 95% CI 1.25–3.15), other determined etiology in TOAST classification (OR 3.00, 95% CI 1.37–6.61), and white matter changes (OR 1.97, 95% CI 1.17–3.33). Conversely, in the case of late recurrent stroke, a significant correlation was noted with the transient ischemic attack (TIA) (OR 2.95, 95% CI 1.52–5.71) and cerebral microbleeds (OR 2.22, 95% CI 1.32–3.75). Additionally, for both early and late recurrent stroke, significant associations were observed with male sex (OR 1.98, 95% CI 1.25–3.15; OR 2.14, 95% CI 1.32–3.48, respectively) and anemia (OR 4.53, 95% CI 2.84–7.20; OR 3.40, 95% CI 2.10–5.51, respectively).

## Discussion

In this study, we focused on understanding the factors influencing stroke recurrence and distinguishing patients experiencing a recurrent stroke from those who experienced a first-ever stroke. Given the dynamic nature of stroke recurrence, we anticipated variations in causative factors over time. Consequently, we categorized our analysis into early and late periods based on a 3-month timeframe.

Our findings indicate that certain factors are closely associated with stroke recurrence within the first 3 months. Patients with diabetes mellitus, other determined etiologies according to the TOAST classification, and white matter changes were particularly prone to early recurrence. By contrast, individuals with TIA and cerebral microbleeds exhibited a higher likelihood of delayed relapse. Regardless of the timing, male sex and anemia were identified as consistent risk factors for recurrence.

Our investigation aligns with previous studies that explored stroke recurrence. Lee et al. discovered that the risk of 1-year stroke recurrence was significantly elevated in men, older adults, and those with a history of ischemic stroke (29). A meta-analysis by Zheng et al. suggested that hypertension, diabetes, atrial fibrillation, and coronary heart disease are associated with a heightened risk of stroke recurrence (30). In a study by Hillen et al., diabetes and atrial fibrillation emerged as significant contributors to outcomes in the first year after the index stroke (31). These risk factors are associated with stroke recurrence, with significant differences. Reviewing multiple prior studies, the most firmly established risk factors linked to stroke recurrence were diabetes and atrial fibrillation (32–35). This is consistent with the current research showing that diabetes is a crucial factor. Regarding atrial fibrillation, our study suggests a tendency for a higher prevalence in patients experiencing recurrent cerebral infarction. However, the results no longer reached statistical significance. This observation could be attributed to the widespread clinical use of NOACs and the heightened early atrial fibrillation detection rate facilitated by tools such as injectable cardiac monitors and long-term Continuous Ambulatory ECG Monitors.

Previous research results on hypertension were more heterogeneous, and several studies reported that hypertension was associated with the highest risk of stroke recurrence. However, few studies reported a significant association with stroke recurrence, which may be related to different criteria and methods for measuring hypertension and issues with antihypertensive management (9,31,32,35,36). Although this study demonstrated a correlation with overall stroke recurrence, no significant correlation was observed with early recurrent stroke.

In our study, a high initial recurrence rate was observed in the category of “other determined etiology” in the TOAST classification, consistent with findings from previous research. Specifically, within this classification, the majority of patients exhibited dissection, Moyamoya disease, and cancer-related coagulopathy, which is consistent with past studies that have identified these conditions as being associated with elevated recurrence rates. For instance, Christian Weimar et al. highlighted the heightened risk of early recurrent stroke following acute ischemic stroke, or TIA, attributed to carotid artery dissection (37). Moreover, a study by Chiu et al. indicated that Moyamoya disease entails a high risk of stroke recurrence, with an 18% recurrence rate within the first year after diagnosis, decreasing to approximately 5% over the subsequent 5 years (38). This trend may be influenced by the gradual development of angiogenesis and collaterals following the initial cerebral infarction. The recurrence rate was higher in patients with cancer-related strokes than in patients with inactive cancer or controls. The estimated one-year stroke recurrence rate in patients with cancer and embolic stroke of undetermined source (ESUS) ranges from 14% to 29%, which is approximately three times higher than that in ESUS patients without cancer (35). White matter changes (WMCs) indicate stroke recurrence up to 5 years after the first ischemic stroke. This shows that white matter change (WMC) can be considered an SVD marker that summarizes the impact of several classical risk factors on small-vessel brain networks (39). We observed that WMC had a particularly strong impact on early recurrent strokes. Conversely, the association between TIA and late recurrent stroke can be attributed to the possibility that symptoms are not observed for an extended duration during the treatment process. The risk factors are effectively managed, and clinicians may opt to discontinue antithrombotic drugs in clinical practice. Considerably, the risk of recurrence in these cases being detected after a long period of time is higher compared to patients with cerebral infarction who consistently adhere to antithrombotic therapy.

Our study has several limitations. First, this was a single-hospital investigation. Inherent in utilizing clinical registry data is a potential bias in patient selection, recording practices, data completeness, and outcome assessment. Therefore, generalizing the findings of this study may be challenging. To bolster the robustness of these results, it is imperative to validate them through prospective multicenter studies with larger sample sizes. Second, a challenge in examining stroke recurrence is the difficulty of identifying such recurrences, particularly if they manifest immediately after the index stroke. Detecting new neurological signs in unconscious, paralyzed, or bedridden patients with a Modified Rankin Scale (mRS) score of 4 or higher is markedly more challenging than in individuals who have recovered. Third, while we categorized stroke recurrence into early (within three months) and late (beyond three months) groups, a more nuanced classification of the late recurrence group could have yielded more detailed insights. Lastly, during the initial 3 months, the loss of follow-up was minimal. However, beyond this period, patients exhibited increased loss of follow-up, potentially introducing bias to our findings.

In conclusion, our study underscores the substantial differences in the causative factors of stroke recurrence, contingent on the timing of recurrence. Effectively mitigating the risk of recurrence in clinical practice necessitates accurately identifying periods when the risk of stroke recurrence is heightened, coupled with stringent control of modifiable risk factors.

## Data Availability

Data are available upon reasonable request to the corresponding author.

## Acknowledgments

None.

## Author contributions

Dr. SH Lee contributed to the study conception and design, data analysis, acquisition of clinical and imaging data, statistical analysis, and manuscript drafting and revision.

Dr. MH Park contributed to the study conception and design, analysis, and interpretation of the imaging and clinical data, manuscript drafting and revision, and study supervision.

Dr. JM Jung contributed to the data analysis and manuscript revision.

Dr. JC Ryu contributed to the study conception and data statistical analysis.

